# Ascending Aortic Calcium Identifies Patients at Risk Despite Normal Myocardial Flow, Perfusion and Coronary Calcium: A 13-Site Cohort Study

**DOI:** 10.64898/2026.10.09.26365170

**Authors:** Clemens P. Spielvogel, Aakash Shanbhag, Shaun Khanna, Tara Zuhair Kassem, Mark Lemley, Jirong Yi, Giselle Ramirez, Paul Kavanagh, Assiata Kamagate, Leandro Slipczuk, Mark I. Travin, Ronny R. Buechel, Robert A. DeKemp, Erick Alexanderson, Isabel Carvajal-Juarez, René R.S. Packard, Mouaz Al-Mallah, Andrew J. Einstein, Wanda Acampa, Stacey Knight, Viet T. Le, Steve Mason, Thomas L. Rosamond, Robert J.H. Miller, Panithaya Chareonthaitawee, Damini Dey, Daniel S. Berman, Marcelo F. Di Carli, Piotr J. Slomka

## Abstract

**Background:** Patients with normal perfusion, normal myocardial flow reserve (MFR) and no coronary artery calcium (CAC) on myocardial perfusion imaging are considered low risk. Yet, adverse events still occur and no routinely reported imaging marker stratifies residual risk. This study aims to determine whether fully automated ascending aortic calcification (AAC) from the attenuation correction CT stratifies risk in these patients.

**Methods:** In the multicenter REFINE-PET registry, AAC was quantified from non-contrast CT using deep learning aortic segmentation and Agatston scoring. Triple-normal was defined as stress total perfusion deficit (TPD) <5%, MFR >2 and CAC 0. The primary endpoint was cardiovascular death, myocardial infarction or heart failure admission, with heart failure admission as key secondary endpoint. Cox models were adjusted for clinical confounders.

**Results:** Of 21,923 patients, 3,580 (16.3%) were triple-normal, of whom 1,322 (36.9%) had AAC (median Agatston 65, IQR 22-212). Agreement with three blinded readers was good to excellent (Intraclass correlation coefficient 0.87-0.90). Over a median follow-up of 52 months (IQR 26-69), 194 primary events occurred. Per standard deviation, AAC was independently associated with the primary endpoint (adjusted HR 1.24, 95% CI 1.09-1.40; p<0.001), heart failure admission (adjusted HR 1.30, 95% CI 1.10-1.55; p=0.003) and all-cause mortality (adjusted HR 1.17, 95% CI 1.04-1.32; p=0.009). AAC improved model fit beyond clinical factors, MFR and TPD (p≤0.009), with modest gains in discrimination (ΔC 0.007-0.009).

**Conclusions:** In patients with normal PET readouts, automated AAC was independently associated with adverse outcomes, most strongly heart failure admission, beyond clinical risk factors, perfusion and flow.

**Clinical Perspective:** *What Is New?:*

- In 3,580 patients with normal perfusion, normal myocardial flow reserve and no coronary artery calcium on PET, more than one in three had ascending aortic calcification on the attenuation correction CT.
- Ascending aortic calcification was independently associated with cardiovascular death, myocardial infarction or heart failure admission (adjusted hazard ratio per SD 1.24, 95% CI 1.09-1.40), most strongly with heart failure admission.
- A fast, fully automated method scored ascending aortic calcification with agreement to expert readers similar to agreement between readers.

*What Are the Clinical Implications?:*

- A normal perfusion PET with normal flow and no coronary calcium does not mean uniformly low risk when the ascending aorta is calcified.
- Because ascending aortic calcification is scored automatically from the CT acquired with every perfusion PET, it could be reported routinely without additional scanning, cost or radiation.
- Patients presumed low risk who have ascending aortic calcification may warrant heart failure-oriented prevention, such as natriuretic peptide screening, pending prospective validation.

## Introduction

Contemporary myocardial perfusion imaging with positron emission tomography (PET) yields a multiparametric readout. Relative perfusion, myocardial flow reserve (MFR) and coronary artery calcium (CAC) from low-dose CT attenuation correction scans are independently prognostic^1,2^. Patients normal across all three are at low short-term risk^3,4^.

However, while cardiovascular event rates after a normal perfusion PET scan are low, they are not zero and because normal studies constitute a large share of clinical volume, the proportion of overall events arising from this group is considerable^5–7^. In this population, residual risk is not evenly distributed. In higher-risk cohorts such as patients with chronic kidney disease, diabetes or advanced age, annualized all-cause mortality after visually normal perfusion imaging has been reported well above 1% and reaches double digits in advanced renal impairment, such that a normal scan identifies lower relative but not low absolute risk^8,9^. These groups represent a growing share of referrals as lower-risk patients are triaged to coronary CT angiography and the prevalence of cardiovascular-kidney-metabolic (CKM) syndrome rises^9,10^. Over the same period, the diagnostic yield of both invasive angiography and noninvasive stress testing has fallen^11–13^ and the phenotype driving events has shifted toward diffuse nonobstructive atherosclerosis and microvascular remodeling. As such, the paradigm on which the negative predictive value of myocardial perfusion imaging rests, the search for flow-limiting epicardial disease, captures a shrinking fraction of the risk carried by the patients now being scanned. Standard PET measures primarily characterize coronary and myocardial disease while not reflecting extracoronary vascular disease. Once perfusion, flow and coronary calcium are all normal, no routinely reported imaging marker remains to separate risk within the group and management defaults to clinical risk factors alone.

Aortic wall calcification addresses this gap. It reflects both intimal atherosclerosis and medial arterial calcification and contributes to aortic stiffening, elevated afterload and left ventricular remodeling, mechanisms that coronary markers capture only indirectly^14,15^. Beyond its impact on hemodynamics, aortic calcification is associated with various cardiovascular and non-cardiovascular outcomes, including stroke, peripheral arterial disease, aneurysm rupture, aortic dissection, reduced coronary blood flow, chronic obstructive pulmonary disease, hip fractures and calcification in other major blood vessels, leading to adverse consequences^16–19^.

Most hybrid imaging examinations, such as perfusion PET/CT, include a non-contrast low-dose CT scan for attenuation correction, covering the ascending aorta. AAC can therefore be assessed opportunistically without additional image acquisition or radiation exposure and no further healthcare system burden. Manual aortic calcium scoring requires an operator to delineate the vessel wall, define segment boundaries and attribute each lesion, a workload that has limited the assessment of vascular calcification in routine reporting despite being present on every cardiac PET/CT. Artificial intelligence (AI) approaches based on automated segmentation models can reduce this workload and enable segment-specific calcium scoring at scale^20–23^.

We therefore developed a fully automated approach for segment-specific aortic calcium quantification from low-dose CT scans and evaluated, in the multicenter REFINE PET registry, whether AAC identifies residual risk in patients whose perfusion, flow and coronary calcium are normal.

## Methods

### Study design and participants

An overview of the study design is shown in **Figure 1**. This study was performed as part of the REFINE PET registry ^1^. We evaluated consecutive patients who underwent myocardial perfusion imaging PET at one of thirteen sites between 2007 and 2020. Patients were excluded if they did not receive a CT scan alongside the PET scan, if the scan did not capture the entire axial range between T5 and T11 vertebrae, or if automated segmentation failed. All eligible registry patients were included and no formal sample size calculation was performed. Patients with total perfusion deficit (TPD) <5%, MFR >2 and no CAC were included for the primary analysis. These patients are referred to as triple-normal patients throughout this manuscript. A secondary analysis was performed for the full cohort.

**Figure 1.**
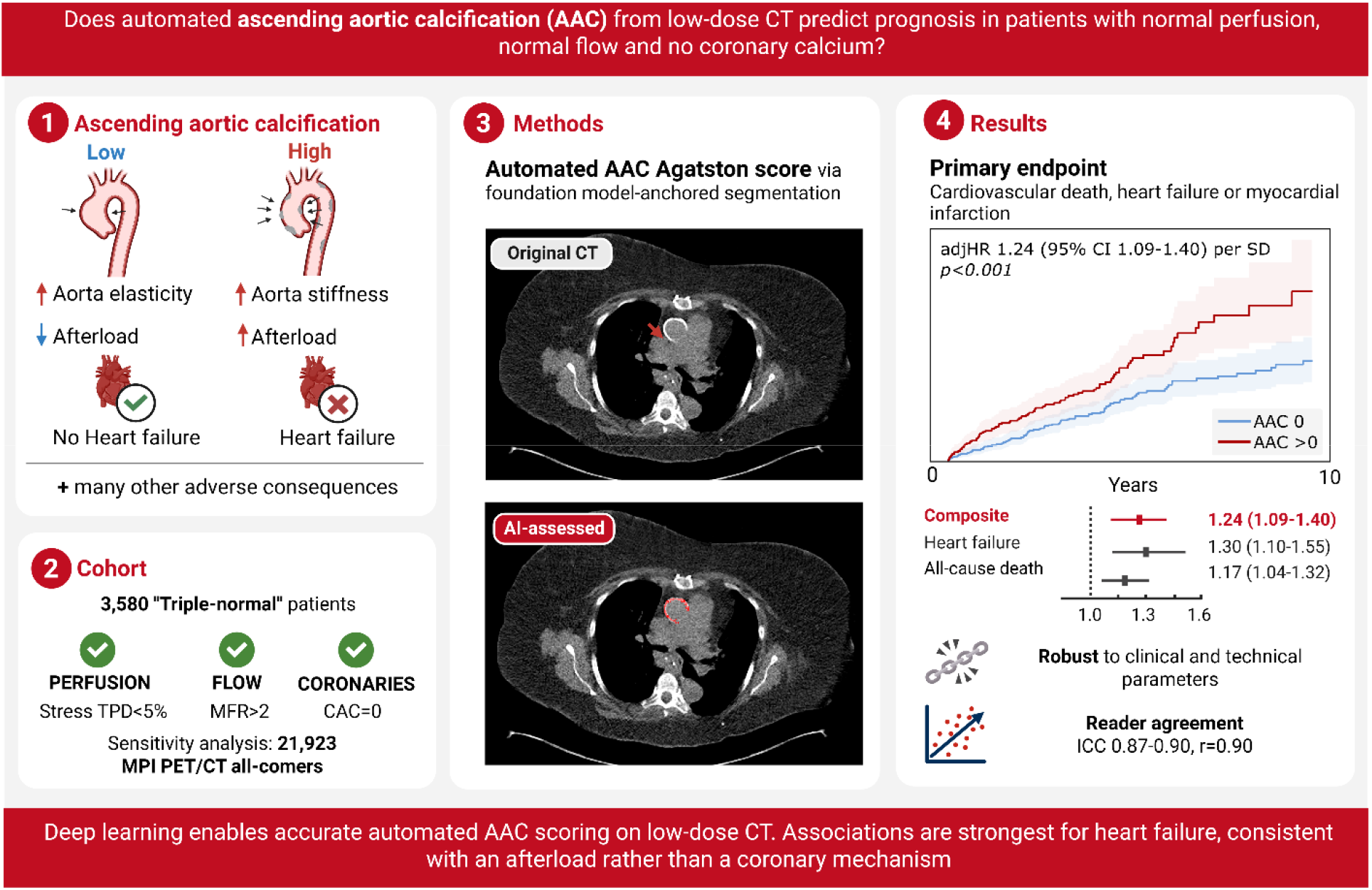
Study overview. AAC=ascending aortic calcification. CI=Confidence interval. HR=hazard ratio. adjHR=adjusted hazard ratio. TPD=total perfusion deficit. MFR=myocardial flow reserve. CAC=coronary artery calcium. ICC=Intraclass correlation coefficient

Sites either obtained written informed consent or the requirement for written informed consent was waived for this retrospective analysis of clinically acquired imaging. This study was conducted in accordance with the Declaration of Helsinki and approved by the institutional review boards at each participating site.

### Endpoints

The primary endpoint was a composite of cardiovascular death, myocardial infarction or heart failure admission with all-cause mortality and heart failure as secondary endpoints. A separate analysis was performed for the individual components of the composite endpoint. All-cause mortality was determined through linkage with the National Death Index. This registry provides the most comprehensive coverage of deaths in the United States and generally identifies more events than the Social Security Administration Death Master File^24^ or institutional electronic health records^25^. Deaths were classified as cardiovascular based on available clinical information and in line with published endpoint definitions^26^. This category covered sudden cardiac death and fatal myocardial infarction, heart failure, stroke, cardiovascular hemorrhage and complications of cardiovascular procedures. Deaths from other cardiovascular conditions, such as pulmonary embolism or peripheral arterial disease, were also included. Cases of incident myocardial infarction were identified by screening electronic health records. An experienced cardiac imaging physician then reviewed each suspected event, drawing on all available clinical findings, laboratory values, imaging studies and invasive coronary angiograms to confirm or reject the diagnosis.

### Imaging and clinical parameters

Hybrid perfusion PET/CT was performed at the corresponding centers and PET/CT images were transferred to the core laboratory at Cedars-Sinai Medical Center. For MFR and TPD calculation, standard clinical routine software within the Cedars-Sinai Cardiac Suite (QPET, Cedars-Sinai Medical Center, Los Angeles) was employed by trained technologists for PET quantification blinded to outcomes. Early dynamic acquisitions were used to measure stress and rest myocardial blood flow, with MFR calculated as stress myocardial blood flow / rest myocardial blood flow. Normal myocardial perfusion was defined as stress TPD <5%, normal flow was defined as MFR >2, and absence of coronary calcium was defined as CAC 0. CAC was measured from low-dose CT scans based on a previously developed approach^22^. Left ventricular ejection fraction was measured from PET. All employed clinical parameters were collected as part of the REFINE PET registry and included age, sex, body mass index, hypertension, diabetes, smoking and estimated glomerular filtration rate (eGFR)^1^.

### Marker quantification

Aortic calcification was quantified fully automatically from the non-contrast CT scan of each rest and stress PET/CT (**Figure 2**). The aorta and all bones were segmented with TotalSegmentator ^27^. To standardize the anatomical coverage across patients, all computation was restricted to the axial region between the T5 and T11 vertebrae. The aorta was subdivided into ascending, arch, and descending thoracic segments by a slice-wise 2D connected-component analysis running from caudal to cranial. The first appearance of at least three consecutive axial slices with two separate lumina defined the onset of the ascending aorta. From there, the descending lumen was tracked across slices as the component with the greatest overlap with the descending lumen below, or the more posterior component in case of a tie. The remaining component was labeled ascending aorta. The first run of at least three consecutive slices returning to a single lumen defined the boundary with the aortic arch.

**Figure 2.**
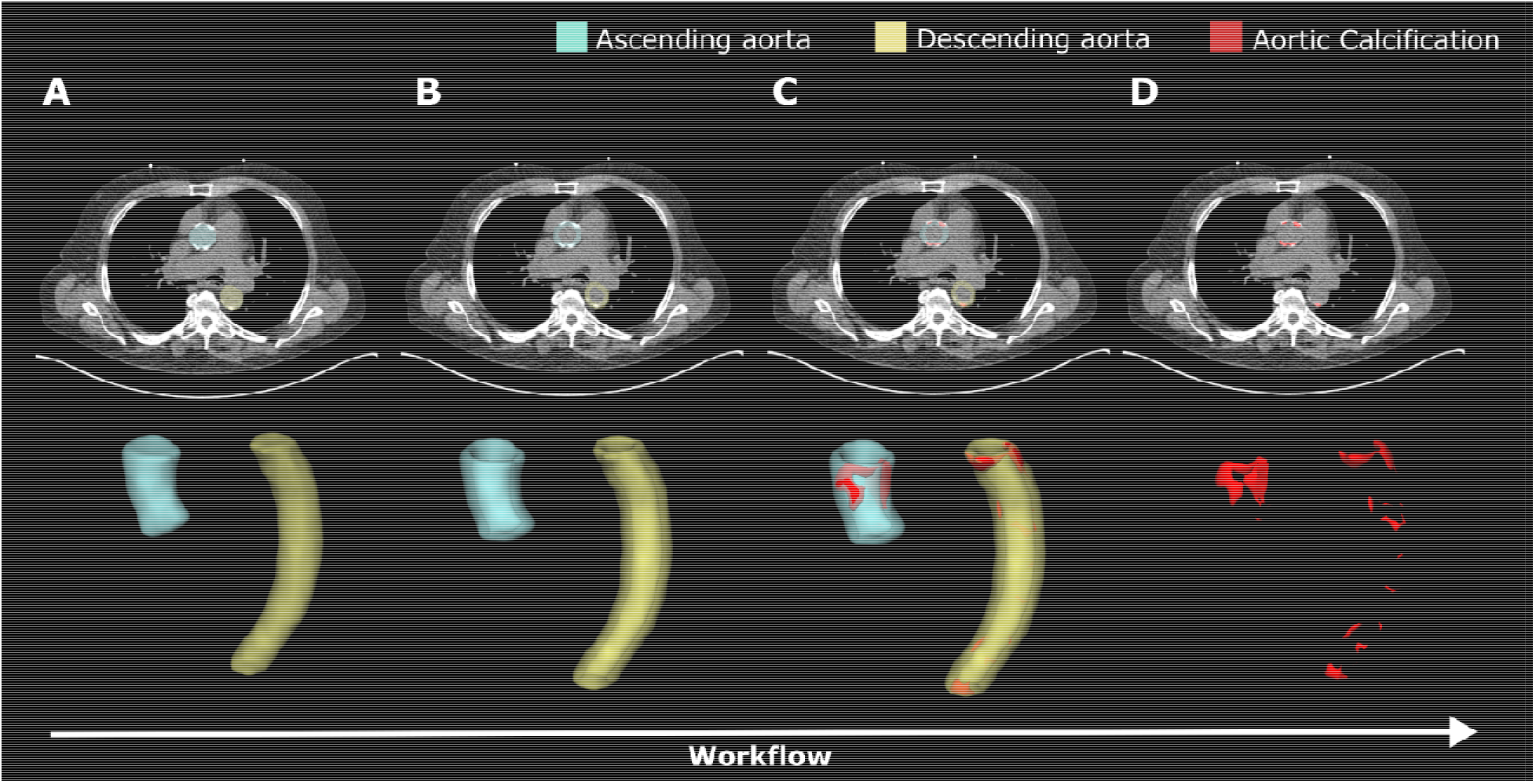
Calcification detection workflow. First, TotalSegmentator aorta segmentations are derived (A). Next, the aorta segmentations are slightly dilated to include the aortic wall and the lumen is removed (B). Subsequently, calcifications are identified in the resulting volume based on Hounsfield unit (>130 HU) thresholding (C). The final calcified lesions are then further processed by computing the Agatston score with lesions contained in the ascending aorta (blue) handled separately from lesions in the descending thoracic aorta (yellow) (D).

For each segment, an aortic wall volume of interest was created by first dilating (expanding) the segmented volume by 3 mm and subsequently subtracting the 5 mm eroded mask to remove the lumen. To avoid collisions between the dilated mask and bone structures, all bone masks were dilated by 4 mm and subtracted from the aortic wall volume. Candidate calcification was defined as ≥130 Hounsfield units. Each lesion was attributed to the segment containing the majority of its voxels, preventing splitting or double counting. To avoid false-positive detection due to noise, lesions below a volume of 0.02 mL were discarded^28,29^. Agatston scores were computed per lesion using standard Agatston scoring with adjustment for slice thickness^30^. The ascending and descending thoracic aortic Agatston scores are the sums of all lesions attributed to the respective segment.

### Subgroup and sensitivity analyses

Measurement reliability was assessed using the paired rest and stress CT acquisitions of the same examination. Since these are separated by minutes and share patient positioning, discordance reflects acquisition and quantification variability rather than biological change. Agreement of continuous AAC was quantified by Pearson correlation. Robustness of the time-to-event analysis was evaluated via multiple sensitivity analyses, including stress versus rest, binarized at the median versus continuous, descending thoracic aorta calcification versus AAC, with relaxed CAC criterion (0-100) versus the strict criterion (CAC 0) and in the full versus the triple-normal cohort.

### Reader study

A total of 244 scans from 122 patients were manually assessed for calcified lesions in the ascending aorta by three experienced (mean 11 years) clinical readers. The readers were blinded to clinical information and to each other. Patients were randomly sampled for each AI-quantified rest AAC range (0, 1-100, 101-400 and >400) from the full cohort. Among the 122 patients, 24 were read by all three readers to determine inter-reader variability. Each reader annotated all calcified lesions using the Cedars-Sinai Cardiac Suite software. For each patient, the readers annotated rest and stress scans separately with a washout duration of at least one day. Inter-reader agreement and reader-deep learning agreement were evaluated via the intraclass correlation coefficient (ICC), specifically ICC (2,1) and using Pearson correlation.

### Statistical analysis

Continuous variables are summarized as mean and standard deviation or median and interquartile ranges (IQR) according to their distribution, which was assessed with the Shapiro-Wilk test. Categorical variables are reported as counts and percentages. Between independent groups, normally distributed continuous variables were compared with the independent samples t-test and non-normally distributed variables with the Mann-Whitney U test. Categorical variables were compared with the Chi-squared test, or Fisher’s exact test where expected cell counts were low.

Time-to-event outcomes were analyzed using cumulative hazard estimates and multivariable Cox proportional hazards models, both unadjusted (u) and adjusted (adj) for prespecified confounders. Confounders for adjustment included age, sex, body mass index, hypertension, diabetes, smoking, eGFR, and left ventricular ejection fraction. The full cohort was additionally adjusted for TPD, MFR and CAC. Confounders were imputed with the median or mode. Endpoints were tested in a fixed hierarchy. The primary endpoint was a composite of cardiovascular death, myocardial infarction or heart failure admission in the triple-normal cohort. Conditional on the primary endpoint being met at a two-sided alpha of 0.05, heart failure admission was tested as the key secondary endpoint at the same alpha. All-cause mortality, individual components of the composite and all analyses in the full cohort, in the CAC 0-100 cohort and using stress rather than rest acquisitions are included as secondary analyses. Fine-Gray models were employed to adjust for competing risks in the primary composite and the secondary heart failure endpoint. Continuous AAC values were log1p-transformed since they are not normally distributed and are zero for a large proportion of patients. Since event and censoring data availability differed slightly across endpoints and because completeness of AAC exposures varied by acquisition (rest vs. stress), the number of patients and events contributing to each Cox model varies marginally across endpoints and predictors. Maximum follow-up time was limited to 10 years. The proportional hazards assumption was tested using Schoenfeld residuals. Restricted cubic spline models with four knots were employed to assess continued associations over the full parameter range. Incremental prognostic value was assessed by comparing nested cause-specific Cox models with and without AAC using the likelihood-ratio test, Harrell’s C (paired bootstrap 95% CI; 1,000 resamples; leave-one-site-out cross-validation) and calibration of cross-validated 5-year risk. Base models comprised the clinical covariates, without and with continuous MFR and stress TPD. All tests were two-sided, and effect estimates were reported with 95% confidence intervals (CIs) and exact p-values. Analyses were performed in Python (version 3.13.13) with the packages SciPy (1.17.1), lifelines (0.30.3), SimpleITK (2.5.5), NumPy (2.4.6) and pandas (2.3.3).

## Results

### Patient characteristics

An overview of the study design is shown in **Figure 1** and the cohort flow diagram in **Supplemental Figure S1**. Of 21,923 patients in the full cohort, 3,580 (16.3%) had normal perfusion (stress TPD <5%), normal flow (MFR >2) and CAC 0, constituting the defined triple-normal cohort. Median age was 60 years (IQR 51-68) and 1,263 patients (35.3%) were male. Hypertension was present in 2,361 patients (65.9%), diabetes in 953 (26.6%) and a history of smoking in 905 (25.3%). AAC was detected in 1,322 patients (36.9%), with a median AAC Agatston score of 65.0 (IQR 22.4-211.9) among those affected. Over a median follow-up of 51.6 months (IQR 26.3-68.8), 194 primary composite events occurred, corresponding to 1.15 events per 100 patient-years.

Patients with any AAC were older (median 65 vs. 58 years; p<0.001) and more frequently had hypertension (71.9% vs. 62.5%; p<0.001) and diabetes (29.0% vs. 25.2%; p=0.030). They had lower renal function (median eGFR 74.0 vs. 81.0 mL/min/1.73 m^2^; p<0.001) and lower MFR within the normal range (median 2.7 vs. 2.9; p<0.001). A smoking history was less frequent in patients with AAC (19.6% vs. 28.6%; p<0.001). Differences in stress TPD (median 1.4% vs. 1.5%; p=0.004) and left ventricular ejection fraction (median 70.8% vs. 70.1%; p=0.011) reached statistical significance but were negligible in magnitude. The clinical characteristics stratified by the presence of AAC are presented in **Table 1**.

**Table 1.** Clinical characteristics. AAC=Ascending aortic calcification. CAC=coronary artery calcium. eGFR=estimated glomerular filtration rate. LVEF=left ventricular ejection fraction. MFR=myocardial flow reserve. SD=standard deviation. TAC=descending thoracic aorta calcification. TPD=total perfusion deficit.

|  |  | Overall | AAC=0 | AAC>0 | P-Value |
| --- | --- | --- | --- | --- | --- |
| Number of patients, n (%) |  | 3580 (100) | 2258 (63.1) | 1322 (36.9) |  |
|  |  | 60.0 | 58.0 | 65.0 |  |
| Age, median [Q1, Q3] |  | [51.0,68.0] | [50.0,66.0] | [55.0,72.0] | <0.001 |
| Male sex, n (%) | yes | 1263 (35.3) | 781 (34.6) | 482 (36.5) | 0.274 |
|  | no | 2317 (64.7) | 1477 (65.4) | 840 (63.5) |  |
|  |  | 31.1 | 30.9 | 31.5 |  |
| Body mass index, median [Q1, Q3] |  | [26.6,37.6] | [26.6,37.2] | [26.5,38.5] | 0.093 |
| eGFR, mL/min/1.73m<sup>2</sup>, median [Q1, Q3] |  | 78.7 | 81.0 | 74.0 |  |
|  |  | [60.0,94.6] | [63.0,96.0] | [60.0,90.9] | <0.001 |
|  |  | 70.4 | 70.1 | 70.8 |  |
| Rest LVEF, median [Q1, Q3] |  | [63.6,76.3] | [63.6,75.7] | [63.6,77.4] | 0.011 |
| MFR, median [Q1, Q3] |  | 2.8 [2.4,3.3] | 2.9 [2.5,3.4] | 2.7 [2.3,3.2] | <0.001 |
| TPD, median [Q1, Q3] |  | 1.5 [0.5,2.8] | 1.5 [0.6,2.8] | 1.4 [0.4,2.7] | 0.004 |
| CAC, mean (SD) |  | 0.0 (0.0) | 0.0 (0.0) | 0.0 (0.0) |  |
|  |  | 0.0 |  | 65.0 |  |
| AAC, median [Q1, Q3] |  | [0.0,30.7] | 0.0 [0.0,0.0] | [22.4,211.9] | <0.001 |
|  |  | 143.9 | 50.2 | 595.1 |  |
| TAC, median [Q1, Q3] |  | [12.2,584.9] | [0.0,218.4] | [184.9,1545.7] | <0.001 |
| Hypertension, n (%) | yes | 2361 (65.9) | 1411 (62.5) | 950 (71.9) | <0.001 |
|  | no | 1179 (32.9) | 808 (35.8) | 371 (28.1) |  |
|  | NA | 40 (1.1) | 39 (1.7) | 1 (0.1) |  |
| Diabetes Mellitus, n (%) | yes | 953 (26.6) | 570 (25.2) | 383 (29.0) | 0.03 |
|  | no | 2625 (73.3) | 1686 (74.7) | 939 (71.0) |  |
|  | NA | 2 (0.1) | 2 (0.1) | 0 (0.0) |  |
| Smoking, n (%) | yes | 905 (25.3) | 646 (28.6) | 259 (19.6) | <0.001 |
|  | no | 2675 (74.7) | 1612 (71.4) | 1063 (80.4) |  |

### Reader study

AI-derived calcified lesions were compared with manual annotations by three experienced clinical readers in 244 scans of 122 patients, of which 24 patients were annotated by all three readers independently. Manual annotation of AAC required a mean of 4.7 minutes per scan. Mean pairwise inter-reader correlation was high (r=0.92, p<0.001), with similar reader-to-deep learning correlation (r=0.90, p<0.001). Agreement between the deep learning approach and the mean of the reader annotations was good on rest acquisitions (ICC 0.87, r=0.88, p<0.001) and excellent on stress acquisitions (ICC 0.90, r=0.91, p<0.001). Agreement was consistent across individual readers, with ICCs of 0.82 to 0.84 on rest and 0.91 to 0.94 on stress. Within-reader agreement between the paired rest and stress acquisitions of the same examination was lower than both (ICC 0.81, range 0.77 to 0.89), indicating that variability attributable to acquisition exceeded variability attributable to the observer. Full results are provided in **Supplemental Table S1**.

### Prognostic associations

Cumulative hazard estimates for the primary composite endpoint stratified by AAC are shown in **Figure 3**. 10-year event rates were 8.1% in patients with no AAC and 13.7% in patients with any AAC. AAC was associated with the primary composite endpoint of cardiovascular death, myocardial infarction or heart failure admission in the unadjusted analysis (unadjusted hazard ratio [uHR] 1.42, 95% CI 1.25-1.60; p<0.001) and remained associated after adjustment for age, sex, body mass index, hypertension, diabetes, smoking, eGFR and left ventricular ejection fraction (adjusted hazard ratio [adjHR] 1.24, 95% CI 1.09-1.40; p<0.001; **Table 2**). The key secondary endpoint, heart failure admission, showed the strongest association of all endpoints tested, both before (uHR 1.63, 95% CI 1.37-1.93; p<0.001) and after adjustment (adjHR 1.30, 95% CI 1.10-1.55; p=0.003). All-cause mortality, as an additional secondary endpoint, was concordantly associated (uHR 1.33, 95% CI 1.19-1.48; p<0.001; adjHR 1.17, 95% CI 1.04-1.32; p=0.009). Accounting for competing risks did not alter these findings, with Fine-Gray subdistribution adjHR of 1.10 (95% CI 1.05-1.14; p<0.001) per standard deviation for the composite (**Supplemental Figure S2**) and 1.17 (95% CI 1.11-1.24; p<0.001) for heart failure admission.

**Figure 3.**
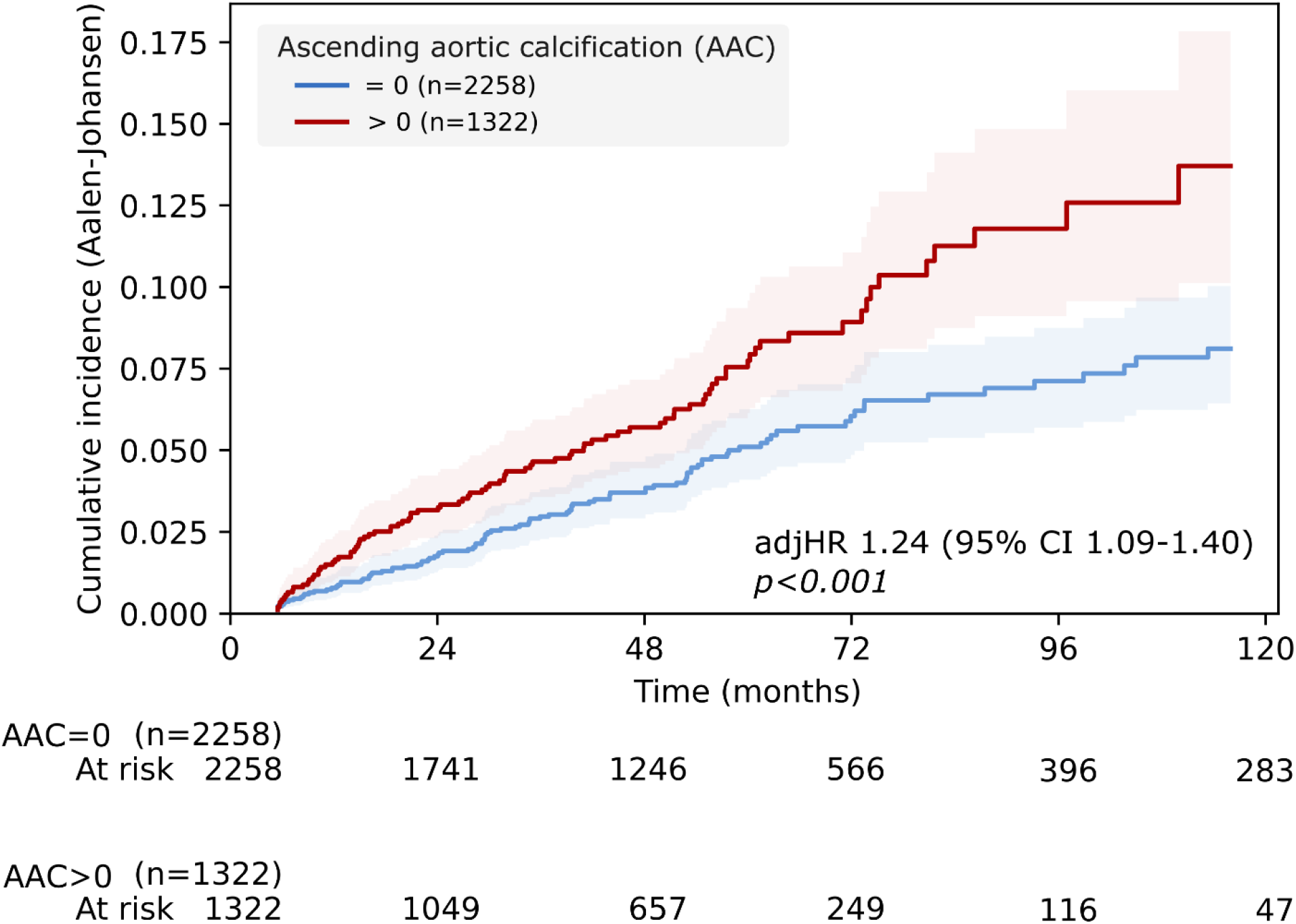
Association of ascending aortic calcification (AAC) Agatston with the primary composite cardiovascular endpoint (heart failure admission or cardiovascular death or myocardial infarction), capped at a follow-up duration of 10 years. adjHR=adjusted hazard ratio. CI=Confidence interval.

**Table 2.** Continuous AAC Cox regression results for the primary and secondary endpoints in the triple-normal cohort. Models were adjusted for age, sex, body mass index, hypertension, diabetes, smoking, estimated glomerular filtration rate (eGFR) and left ventricular ejection fraction. adjHR=adjusted hazard ratio. CI=Confidence interval. uHR=unadjusted hazard ratio.

| Endpoint | uHR (95% CI) | p value<br>(unadjusted) | AdjHR (95% CI) | p value<br>(adjusted) |
| --- | --- | --- | --- | --- |
| Composite | 1.42 (1.25-1.60) | <0.001 | 1.24 (1.09-1.40) | <0.001 |
| Heart failure admission | 1.63 (1.37-1.93) | <0.001 | 1.30 (1.10-1.55) | 0.003 |
| Death | 1.33 (1.19-1.48) | <0.001 | 1.17 (1.04-1.32) | 0.009 |

Restricted cubic spline analysis demonstrated a significant monotonic increase in hazard across the observed AAC range without evidence of a threshold effect (**Figure 4**, p=0.005). Risk was already elevated at low absolute Agatston scores, with the adjusted hazard of AAC increasing at any Agatston units above zero. An adjHR of 2.00 compared to AAC>0 was reached at approximately AAC>1000.

**Figure 4.**
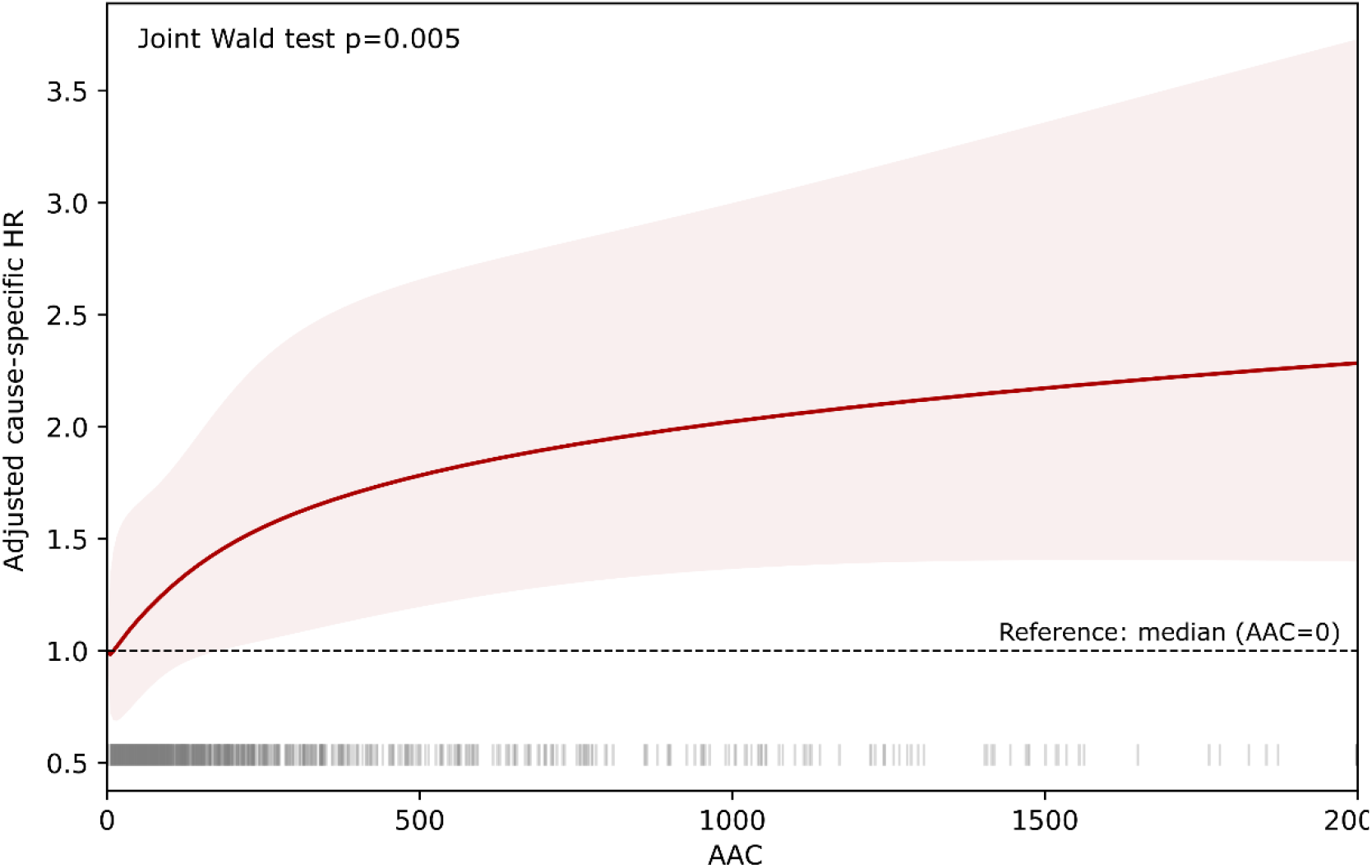
The adjusted hazard ratio for the primary composite cardiovascular endpoint continuously increases with higher AAC Agatston scores. Rug lines (vertical gray lines above the x axis) indicate the AAC distribution. AAC=ascending aortic calcification. HR=Hazard ratio.

Components of the primary composite are shown in **Supplemental Figure S3**. Of 194 primary events, 95 (49%) were heart failure admissions, 65 (33%) myocardial infarctions and 34 (18%) cardiovascular deaths without previous heart failure admission or myocardial infarction during follow-up. Adjusted HRs per standard deviation of AAC were 1.30 (95% CI 1.10-1.55) for heart failure admission, 1.15 (95% CI 0.84-1.57) for cardiovascular death and 1.07 (95% CI 0.84-1.38) for myocardial infarction.

Adding AAC to the clinical model improved model fit for the primary endpoint (likelihood-ratio χ^2^ 9.3; p=0.002) and for heart failure admission (χ^2^ 8.9; p=0.003), including when MFR and stress TPD were added to the base model (p=0.004 and p=0.009). Discrimination increased modestly (primary endpoint C 0.685 to 0.693, ΔC 0.007, 95% CI −0.001 to 0.024; heart failure admission C 0.801 to 0.810, ΔC 0.009, 95% CI −0.001 to 0.027) and was consistent under leave-one-site-out cross-validation. Calibration of 5-year risk was preserved (calibration slope 0.88 to 0.89, observed/expected ratio 0.97 to 1.00; **Supplemental Table S2**).

### Subgroup and sensitivity analyses

Sensitivity analyses are summarized in **Supplemental Table S3**. Rest and stress AAC were strongly correlated (r=0.83; p<0.001) and substituting stress for rest acquisitions left the primary association essentially unchanged (uHR 1.45, 95% CI 1.28-1.64; p<0.001; adjHR 1.26, 95% CI 1.11-1.43; p<0.001).

The association was preserved when the full cohort was analyzed (uHR 1.48, 95% CI 1.43-1.53; p<0.001; adjHR 1.11, 95% CI 1.07-1.15; p<0.001). The adjusted point estimate per standard deviation was smaller in the full cohort than in the triple-normal cohort (1.11 vs. 1.24) while being estimated with substantially greater precision, consistent with AAC carrying the most incremental information where the coronary readout is uninformative. The corresponding analysis in the CAC 0-100 cohort demonstrated a significant association with the primary endpoint, unchanged compared to the stricter (CAC 0) definition of triple-normal (uHR 1.38, 95% CI 1.25-1.53; p<0.001; adjHR 1.23, 95% CI 1.11-1.37; p<0.001).

Calcification of the descending thoracic aorta, scored as a separate segment, was likewise associated with the primary endpoint (uHR 1.37, 95% CI 1.19-1.59; p<0.001; adjHR 1.19, 95% CI 1.03-1.39; p=0.022) and correlated only moderately with AAC (r=0.52, p<0.001). Dichotomizing AAC at 0, which also represented the median, yielded a consistent effect (uHR 1.76, 95% CI 1.34-2.33; p<0.001; adjHR 1.44, 95% CI 1.08-1.91; p=0.014).

## Discussion

In this multicenter study of 21,923 consecutive patients undergoing perfusion PET, fully automated AAC was independently associated with adverse outcomes among the 3,580 patients with normal perfusion, normal myocardial flow and no coronary calcium. AAC was associated with the primary composite endpoint (adjHR 1.24 per SD, 95% CI 1.09-1.40), with heart failure admission as the key secondary endpoint (adjHR 1.30, 95% CI 1.10-1.55) and with all-cause mortality (adjHR 1.17, 95% CI 1.04-1.32). The associations were independent of clinical risk factors and of perfusion as well as flow within the normal range, although the gain in discrimination was modest, as expected when a single marker is added to an established risk model^31–33^.

The association with heart failure admission shows hemodynamic contribution through increased aortic stiffness and ventricular afterload rather than a purely atherosclerotic pathway. The ascending aorta is the principal contributor to the Windkessel reservoir, so stiffening at this level raises the pressure the ventricle must generate during early ejection, when instantaneous wall stress peaks^14^. Proximal stiffening increases early systolic afterload, while stiffening along the remainder of the aorta amplifies wave reflection and shifts reflected load earlier into systole, adding a late systolic afterload component repeatedly linked to concentric remodeling, impaired relaxation and incident heart failure in community cohorts^14,15^. Aortic wall calcification serves as an indirect marker of these vascular changes and can be quantified opportunistically from routinely acquired CT images.

Left ventricular ejection fraction was preserved in both groups and differed by less than one percentage point (median 70.8% versus 70.1%). The residual risk identified by AAC is therefore invisible to the only measure of ventricular performance the PET study already reports. This is consistent with a trajectory toward heart failure with preserved ejection fraction, in which ejection fraction is normal by definition. Ejection fraction alone is therefore an inadequate screen in this population.

In the Multi-Ethnic Study of Atherosclerosis, thoracic aortic calcium predicted coronary heart disease and cardiovascular events, although its incremental value over coronary calcium was modest ^34^. Razavi et al. recently showed across the full spectrum of coronary calcium burden that thoracic aortic calcium remained associated with incident cardiovascular disease and all-cause mortality, with the strongest associations in participants with CAC 0^35^. This study extends these observations to a clinically referred population with normal perfusion, preserved MFR and no CAC, using segment-specific quantification of aortic calcium. First, MFR forms part of the negative-test definition, so the residual risk identified here persists in patients without impairment of global myocardial flow reserve, which a calcium-only cohort cannot do. Second, the aorta was scored in separate segments rather than as a single territory. Third, the population consists of symptomatic patients referred for testing rather than asymptomatic community volunteers, leading to different absolute risk and clinical decision-making.

The workload of manual scoring has kept extracoronary calcium out of routine reporting of low-dose CT images. In our reader study, annotation required a mean of 4.7 minutes per scan, which for the triple-normal cohort alone would amount to several hundred reader-hours. Anchoring quantification on a foundation segmentation model removes this bottleneck while avoiding the requirement of re-training an artificial intelligence model^27^. Agreement between the automated AAC scores and expert readers was good to excellent (ICC 0.87-0.90) and was equal to that of inter-reader agreement. The same approach of deriving quantitative imaging biomarkers by anchoring simple modifications on general-purpose segmentation models has recently been applied to perivascular adipose tissue and tracer uptake in cardiac amyloidosis^23,36^.

Normal studies dominate cardiac PET volume overall^37,38^. Triple-normal patients accounted for 16.3% of consecutive referrals in the employed real-world multicenter registry. These patients may currently be reassured and discharged without targeted follow-up examinations. As such, even a low event rate translates into a considerable absolute number of events arising from a group that receives no further clinical attention. CT scans are acquired in every PET/CT examination, including oncological and other non-cardiac exams. Opportunistic AAC quantification requires no additional image acquisition, contrast administration or radiation exposure and could be incorporated into PET/CT reporting as an additional quantitative measure This would be an extension of existing guidance, which already recommends opportunistic evaluation of coronary calcium and incidental findings on hybrid nuclear CT attenuation maps^39^.

If confirmed prospectively, elevated AAC could prompt targeted heart failure work-up. The predominance of heart failure admissions suggests that preventive attention in triple-normal patients with AAC should extend to heart failure risk, in addition to atherosclerotic risk factor management, for example through natriuretic peptide-based screening as recommended for patients at risk of heart failure^40^.

Women accounted for 64.7% of the triple-normal cohort, reflecting the known enrichment of normal perfusion studies and of CAC 0 among women^41^, who simultaneously carry a disproportionate burden of heart failure with preserved ejection fraction and of non-obstructive coronary disease^42,43^.

This study has several limitations. Our data are consistent with, but do not establish, a stiffening-mediated pathway. We had no direct measure of aortic stiffness such as pulse wave velocity or augmentation index. Calcification extent is a surrogate of stiffening rather than a quantification of it. The analysis is observational, so residual confounding by unmeasured factors such as mineral metabolism, systemic inflammation, duration of hypertension or visceral adiposity, which BMI captures incompletely^44^, cannot be excluded. The aortic arch could not be scored because it is frequently outside of the field of view. Absolute risk in this population remains low. The gain in discrimination was modest. A substantial share of events occurred in patients without AAC. This is expected, given that AAC was absent in 63% of the cohort and that events in this population also arise through non-calcific pathways such as hypertensive or metabolic heart disease. Absence of AAC therefore does not exclude risk. The cohort included thirteen sites across North America and Europe with differing scanners and protocols. Because AAC is an anatomical marker independent of the perfusion tracer, it can in principle be derived from any low-dose CT covering the ascending aorta, including SPECT/CT and oncological PET/CT. However, we did not evaluate AAC in the context of these imaging modalities.

## Conclusions

Even when perfusion, flow and coronary calcium are all normal, calcification of the ascending aorta signals residual risk, particularly for heart failure. Scored fully automatically from the attenuation-corrected CT without additional scanning, cost or radiation, it could identify patients presumed low risk who merit heart failure-oriented prevention.

## Data Availability

The data that support the findings of this study are available from the corresponding author upon reasonable request, subject to the data use agreements governing the REFINE PET registry and to institutional review board approval at the contributing sites.

## Acknowledgements

Biorender: Figure 1; Flaticon (SBTS2018, Magnific): Figure 1

## Data access

CPS had full access to all the data and takes responsibility for its integrity and the data analysis.

## Author Contributions

CPS conceived the study. CPS and PJS planned the study. CPS performed the analysis. ML, GR, PK, AK, LS, MIT, EA, RB, RDK, ICJ, RRSP, MAM, AJE, WA, SK, VTL, SM, RJHM, PC, TLR, MFDC, DD, DSB and PJS contributed to data collection. SK, ML and TZK performed the clinical reference reading. CPS and ML set up the reading study and provided technical guidance for the clinical readers. DSB and RJHM provided clinical consulting. JY, AS and PJS provided technical feedback. CPS and AS wrote the associated code. CPS created all figures and wrote the initial draft. All co-authors provided feedback and agreed to the publication of the study.

## Nonstandard Abbreviations and Acronyms

AAC: ascending aortic calcification
adjHR: adjusted hazard ratio
CAC: coronary artery calcium
eGFR: estimated glomerular filtration rate
ICC: intraclass correlation coefficient
MFR: myocardial flow reserve
TPD: total perfusion deficit
uHR: unadjusted hazard ratio

## References

1. Ramirez G, Lemley M, Shanbhag A, Kwiecinski J, Miller RJH, Kavanagh PB, Liang JX, Dey D, Slipczuk L, Travin MI, et al. The REgistry of Flow and Perfusion Imaging for Artificial Intelligence with positron emission tomography (REFINE PET):Rationale and design. J Nucl Cardiol. 2025;52:102449. doi: 10.1016/j.nuclcard.2025.102449

2. Bateman TM, Heller GV, Beanlands R, Calnon DA, Case J, deKemp R, DePuey EG, Di Carli M, Guler EC, Murthy VL, et al. Practical Guide for Interpreting and Reporting Cardiac PET Measurements of Myocardial Blood Flow: An Information Statement from the American Society of Nuclear Cardiology, and the Society of Nuclear Medicine and Molecular Imaging. J Nucl Med. 2021;62:1599–1615. doi: 10.2967/jnumed.121.261989

3. Aljizeeri A, Ahmed AI, Alfaris MA, Ahmed D, Farea J, Elneama A, Suliman I, Alharthi M, Ahmed A, Alsaileek A, et al. Myocardial Flow Reserve and Coronary Calcification in Prognosis of Patients With Suspected Coronary Artery Disease. JACC Cardiovasc Imaging. 2021;14:2443–2452. doi: 10.1016/j.jcmg.2021.01.024

4. Patel KK, Peri-Okonny PA, Qarajeh R, Patel FS, Sperry BW, McGhie AI, Thompson RC, Kennedy KF, Chan PS, Spertus JA, et al. Prognostic Relationship Between Coronary Artery Calcium Score, Perfusion Defects, and Myocardial Blood Flow Reserve in Patients With Suspected Coronary Artery Disease. Circ Cardiovasc Imaging. 2022;15:e012599. doi: 10.1161/CIRCIMAGING.121.012599

5. Dorbala S, Di Carli MF. Cardiac PET perfusion: prognosis, risk stratification, and clinical management. Semin Nucl Med. 2014;44:344–357. doi: 10.1053/j.semnuclmed.2014.05.003

6. Dorbala S, Di Carli MF, Beanlands RS, Merhige ME, Williams BA, Veledar E, Chow BJ, Min JK, Pencina MJ, Berman DS, et al. Prognostic value of stress myocardial perfusion positron emission tomography: results from a multicenter observational registry. J Am Coll Cardiol. 2013;61:176–184. doi: 10.1016/j.jacc.2012.09.043

7. Skali H, Di Carli MF, Blankstein R, Chow BJ, Beanlands RS, Berman DS, Germano G, Min JK, Merhige M, Williams B, et al. Stress Myocardial Perfusion PET Provides Incremental Risk Prediction in Patients with and Patients without Diabetes. Radiol Cardiothorac Imaging. 2019;1:e180018. doi: 10.1148/ryct.2019180018

8. Al-Mallah MH, Hachamovitch R, Dorbala S, Di Carli MF. Incremental prognostic value of myocardial perfusion imaging in patients referred to stress single-photon emission computed tomography with renal dysfunction. Circ Cardiovasc Imaging. 2009;2:429–436. doi: 10.1161/CIRCIMAGING.108.831164

9. Di Carli MF. Future of Radionuclide Myocardial Perfusion Imaging: Transitioning from SPECT to PET. J Nucl Med. 2023;64:3S–10S. doi: 10.2967/jnumed.122.264864

10. Rozanski A, Gransar H, Miller R, Slomka P, Han D, Hayes S, Friedman J, Berman D. Temporal trends in the clinical utilization of cardiac imaging tests: 2008-2024. J Cardiovasc Comput Tomogr. 2026;20:347–353. doi: 10.1016/j.jcct.2026.04.006

11. Patel MR, Peterson ED, Dai D, Brennan JM, Redberg RF, Anderson HV, Brindis RG, Douglas PS. Low diagnostic yield of elective coronary angiography. N Engl J Med. 2010;362:886–895. doi: 10.1056/NEJMoa0907272

12. Rozanski A, Gransar H, Hayes SW, Min J, Friedman JD, Thomson LE, Berman DS. Temporal trends in the frequency of inducible myocardial ischemia during cardiac stress testing: 1991 to 2009. J Am Coll Cardiol. 2013;61:1054–1065. doi: 10.1016/j.jacc.2012.11.056

13. Taqueti VR, Di Carli MF. Coronary Microvascular Disease Pathogenic Mechanisms and Therapeutic Options: JACC State-of-the-Art Review. J Am Coll Cardiol. 2018;72:2625–2641. doi: 10.1016/j.jacc.2018.09.042

14. Chirinos JA, Segers P, Hughes T, Townsend R. Large-Artery Stiffness in Health and Disease: JACC State-of-the-Art Review. J Am Coll Cardiol. 2019;74:1237–1263. doi: 10.1016/j.jacc.2019.07.012

15. Tsao CW, Lyass A, Larson MG, Levy D, Hamburg NM, Vita JA, Benjamin EJ, Mitchell GF, Vasan RS. Relation of Central Arterial Stiffness to Incident Heart Failure in the Community. J Am Heart Assoc. 2015;4. doi: 10.1161/JAHA.115.002189

16. Thomas IC, Thompson CA, Yang M, Allison MA, Forbang NI, Michos ED, McClelland RL, Budoff MJ, Criqui MH. Thoracic Aorta Calcification and Noncardiovascular Disease-Related Mortality. Arterioscler Thromb Vasc Biol. 2018;38:1926–1932. doi: 10.1161/ATVBAHA.118.310850

17. Iribarren C, Sidney S, Sternfeld B, Browner WS. Calcification of the aortic arch: risk factors and association with coronary heart disease, stroke, and peripheral vascular disease. JAMA. 2000;283:2810–2815. doi: 10.1001/jama.283.21.2810

18. Petersen CS, Karon Z, Obel LM, Hasific S, Mejldal A, Krasniqi L, Schaeffer M, Steffensen FH, Frost L, Lambrechtsen J, et al. Aortic and Iliac Calcifications as Predictors of Aortic Dissection, Aneurysm Rupture, and Peripheral Vascular Disease: A Prospective Cohort Study from the DANCAVAS Trials. Circulation. 2026;153:1026–1037. doi: 10.1161/CIRCULATIONAHA.125.076240

19. Ribeiro LC, Almeida M, Malheiro J, Silva F, Nunes-Carneiro D, Martins S, Pedroso S, Silva-Ramos M. Association of the Calcification Score of the Abdominal Aorta, Common Iliac, and Renal Arteries with Outcomes in Living Kidney Donors. J Clin Med. 2023;12. doi: 10.3390/jcm12093339

20. Miller RJ, Shanbhag A, Shahi K, Bosley D, Litwin L, White JA, Jimenez-Zepeda V, Dey D, Berman DS, Fine NM, et al. Artificial intelligence-driven longitudinal quantification of technetium pyrophosphate uptake in cardiac amyloidosis: Correlation with multimodality imaging and outcomes. J Nucl Cardiol. 2026;56:102573. doi: 10.1016/j.nuclcard.2025.102573

21. Miller RJH, Yi J, Shanbhag A, Marcinkiewicz A, Patel KK, Lemley M, Ramirez G, Geers J, Chareonthaitawee P, Wopperer S, et al. Deep learning-quantified body composition from positron emission tomography/computed tomography and cardiovascular outcomes: a multicentre study. Eur Heart J. 2025;46:2336–2347. doi: 10.1093/eurheartj/ehaf131

22. Pieszko K, Shanbhag A, Killekar A, Miller RJH, Lemley M, Otaki Y, Singh A, Kwiecinski J, Gransar H, Van Kriekinge SD, et al. Deep Learning of Coronary Calcium Scores From PET/CT Attenuation Maps Accurately Predicts Adverse Cardiovascular Events. JACC Cardiovasc Imaging. 2023;16:675–687. doi: 10.1016/j.jcmg.2022.06.006

23. Spielvogel CP, Kofler M, Jiang Z, Ning J, Yu J, Haberl D, Kronberger C, Poledniczek M, Schmid LM, Kersting D, et al. Impact of disease-modifying therapy on [(99m)Tc]Tc-DPD SPECT/CT markers in transthyretin cardiac amyloidosis enabled by artificial intelligence. Eur J Nucl Med Mol Imaging. 2026;53:3310–3324. doi: 10.1007/s00259-025-07673-6

24. Navar AM, Peterson ED, Steen DL, Wojdyla DM, Sanchez RJ, Khan I, Song X, Gold ME, Pencina MJ. Evaluation of Mortality Data From the Social Security Administration Death Master File for Clinical Research. JAMA Cardiol. 2019;4:375–379. doi: 10.1001/jamacardio.2019.0198

25. Ter-Minassian M, Basra SS, Watson ES, Derus AJ, Horberg MA. Validation of US CDC National Death Index mortality data, focusing on differences in race and ethnicity. BMJ Health Care Inform. 2023;30. doi: 10.1136/bmjhci-2023-100737

26. Hicks KA, Mahaffey KW, Mehran R, Nissen SE, Wiviott SD, Dunn B, Solomon SD, Marler JR, Teerlink JR, Farb A, et al. 2017 Cardiovascular and Stroke Endpoint Definitions for Clinical Trials. Circulation. 2018;137:961–972. doi: 10.1161/CIRCULATIONAHA.117.033502

27. Wasserthal J, Breit HC, Meyer MT, Pradella M, Hinck D, Sauter AW, Heye T, Boll DT, Cyriac J, Yang S, et al. TotalSegmentator: Robust Segmentation of 104 Anatomic Structures in CT Images. Radiol Artif Intell. 2023;5:e230024. doi: 10.1148/ryai.230024

28. Isgum I, de Vos BD, Wolterink JM, Dey D, Berman DS, Rubeaux M, Leiner T, Slomka PJ. Erratum to: Automatic determination of cardiovascular risk by CT attenuation correction maps in Rb-82 PET/CT. J Nucl Cardiol. 2018;25:2143. doi: 10.1007/s12350-017-0946-4

29. Bielak LF, Kaufmann RB, Moll PP, McCollough CH, Schwartz RS, Sheedy PF, 2nd. Small lesions in the heart identified at electron beam CT: calcification or noise? Radiology. 1994;192:631–636. doi: 10.1148/radiology.192.3.8058926

30. Agatston AS, Janowitz WR, Hildner FJ, Zusmer NR, Viamonte M, Jr., Detrano R. Quantification of coronary artery calcium using ultrafast computed tomography. J Am Coll Cardiol. 1990;15:827–832. doi: 10.1016/0735-1097(90)90282-t

31. Pepe MS, Janes H, Longton G, Leisenring W, Newcomb P. Limitations of the odds ratio in gauging the performance of a diagnostic, prognostic, or screening marker. Am J Epidemiol. 2004;159:882–890. doi: 10.1093/aje/kwh101

32. Khanna S, Ho W, Takeuchi F, Soh CH, Bhat A, Lin A, Patel A, Thomas L, Marwick TH, Nerlekar N, et al. Regional fat distribution as a determinant of mortality in populations with left ventricular systolic dysfunction: challenging the obesity paradox. Eur J Prev Cardiol. 2026. doi: 10.1093/eurjpc/zwag310

33. Cook NR. Use and misuse of the receiver operating characteristic curve in risk prediction. Circulation. 2007;115:928–935. doi: 10.1161/CIRCULATIONAHA.106.672402

34. Budoff MJ, Nasir K, Katz R, Takasu J, Carr JJ, Wong ND, Allison M, Lima JA, Detrano R, Blumenthal RS, et al. Thoracic aortic calcification and coronary heart disease events: the multi-ethnic study of atherosclerosis (MESA). Atherosclerosis. 2011;215:196–202. doi: 10.1016/j.atherosclerosis.2010.11.017

35. Razavi AC, Dzaye O, Cainzos-Achirica M, Dardari Z, Van Assen M, Quyyumi AA, Nasir K, Carr JJ, Budoff MJ, Blumenthal RS, et al. Association of thoracic aortic calcium with incident cardiovascular disease and all-cause mortality across the spectrum of coronary artery calcium burden. Am J Prev Cardiol. 2025;21:100916. doi: 10.1016/j.ajpc.2024.100916

36. Goudot G, Abohashem S, Osborne MT, Aldosoky W, Ahmad TZ, Lu MT, Foldyna B, Tawakol A. Periaortic Fat Attenuation on Nongated Noncontrast Chest CT Images to Assess Changes in Arterial Inflammation: Impact of Atorvastatin. Circ Cardiovasc Imaging. 2025;18:e017248. doi: 10.1161/CIRCIMAGING.124.017248

37. Ramirez G, Builoff V, Miller RJH, Lemley M, Carvajal-Juarez I, Alexanderson E, Rosamond TL, Song N, Travin MI, Slipczuk L, et al. Multicenter Evaluation of Myocardial Flow Reserve as a Prognostic Marker for Mortality in N-Ammonia PET Myocardial Perfusion Imaging. Circ-Cardiovasc Imag. 2026;19. doi: ARTN e018729 10.1161/CIRCIMAGING.125.018729

38. Lee SN, Malhotra P, Miller RJH, Gransar H, Hayes SW, Friedman JD, Thomson LEJ, Rozanski A, Slomka PJ, Han D, et al. Independent prognostic significance of myocardial flow reserve over coronary artery calcium, myocardial perfusion, and clinical variables in patients without known coronary artery disease, according to diabetes status. J Nucl Cardiol. 2025;47:102165. doi: 10.1016/j.nuclcard.2025.102165

39. Al-Mallah MH, Bateman TM, Branch KR, Crean A, Gingold EL, Thompson RC, McKenney SE, Miller EJ, Murthy VL, Nieman K, et al. 2022 ASNC/AAPM/SCCT/SNMMI guideline for the use of CT in hybrid nuclear/CT cardiac imaging. J Nucl Cardiol. 2022;29:3491–3535. doi: 10.1007/s12350-022-03089-z

40. Heidenreich PA, Bozkurt B, Aguilar D, Allen LA, Byun JJ, Colvin MM, Deswal A, Drazner MH, Dunlay SM, Evers LR, et al. 2022 AHA/ACC/HFSA Guideline for the Management of Heart Failure: A Report of the American College of Cardiology/American Heart Association Joint Committee on Clinical Practice Guidelines. Circulation. 2022;145:e895–e1032. doi: 10.1161/CIR.0000000000001063

41. McClelland RL, Chung H, Detrano R, Post W, Kronmal RA. Distribution of coronary artery calcium by race, gender, and age: results from the Multi-Ethnic Study of Atherosclerosis (MESA). Circulation. 2006;113:30–37. doi: 10.1161/CIRCULATIONAHA.105.580696

42. Beale AL, Meyer P, Marwick TH, Lam CSP, Kaye DM. Sex Differences in Cardiovascular Pathophysiology: Why Women Are Overrepresented in Heart Failure With Preserved Ejection Fraction. Circulation. 2018;138:198–205. doi: 10.1161/CIRCULATIONAHA.118.034271

43. Waheed N, Elias-Smale S, Malas W, Maas AH, Sedlak TL, Tremmel J, Mehta PK. Sex differences in non-obstructive coronary artery disease. Cardiovasc Res. 2020;116:829–840. doi: 10.1093/cvr/cvaa001

44. Khanna S, Mann G, Bhat A, Lin A, Patel A, Nicholls SJ, Sindone A, Dey D, Bhatt DL, Arnott C, et al. Role of systemic and epicardial adipose tissue in cardiometabolic disease. Nat Rev Cardiol. 2026. doi: 10.1038/s41569-026-01304-9

