## Supplemental Material for "Ascending Aortic Calcium Identifies Patients at Risk Despite Normal Myocardial Flow, Perfusion and Coronary Calcium: A 13-Site Cohort Study"

**Supplement**

**Table S1.** Inter-rater variability and agreement with deep learning pipeline. AI=Artificial intelligence (automated approach). ICC=Intraclass correlation coefficient.

| **Annotator 1** | **Annotator 2** | **Scan** | **ICC** | **r** | **p value** |
| --- | --- | --- | --- | --- | --- |
| Reader 3 | AI | Rest | 0.84 | 0.85 | <0.001 |
| Reader 3 | AI | Stress | 0.91 | 0.92 | <0.001 |
| Reader 2 | AI | Rest | 0.83 | 0.85 | <0.001 |
| Reader 2 | AI | Stress | 0.94 | 0.95 | <0.001 |
| Reader 1 | AI | Rest | 0.82 | 0.86 | <0.001 |
| Reader 1 | AI | Stress | 0.94 | 0.95 | <0.001 |
| Mean | AI | Rest | 0.87 | 0.88 | <0.001 |
| Mean | AI | Stress | 0.90 | 0.91 | <0.001 |
| **Mean** | **AI** | **Rest and stress** | **0.88** | **0.90** | <0.001 |
| Reader 3 | Reader 2 | Rest | 0.84 | 0.90 | <0.001 |
| Reader 3 | Reader 2 | Stress | 0.95 | 0.97 | <0.001 |
| Reader 3 | Reader 1 | Rest | 0.64 | 0.76 | <0.001 |
| Reader 3 | Reader 1 | Stress | 0.98 | 0.99 | <0.001 |
| Reader 2 | Reader 1 | Rest | 0.90 | 0.93 | <0.001 |
| Reader 2 | Reader 1 | Stress | 0.98 | 0.98 | <0.001 |
| **Mean** | **Mean** | **Rest and stress** | **0.88** | **0.92** | <0.001 |
| Reader 3 | Reader 3 | Rest vs stress | 0.77 | 0.77 | <0.001 |
| Reader 2 | Reader 2 | Rest vs stress | 0.89 | 0.89 | <0.001 |
| Reader 1 | Reader 1 | Rest vs stress | 0.78 | 0.82 | <0.001 |
| **Mean** | **Mean** | **Rest vs stress** | **0.81** | **0.83** | <0.001 |

**Table S2.** Incremental prognostic value of AAC beyond clinical and PET parameters in the triple-normal cohort. Nested cause-specific Cox models with and without continuous AAC. LR χ² and P from the likelihood-ratio test of the nested models (1 df). AAC=ascending aortic calcification; CI=confidence interval; LR=likelihood ratio; MFR=myocardial flow reserve; O/E=observed-to-expected ratio; TPD=total perfusion deficit.

| **Endpoint** | **Base model** | **LR χ²** | **P** | **C base** | **C base + AAC** | **ΔC**  **(95% CI)** | **ΔC cross-validated (95% CI)** | **Calibration slope, base / + AAC** | **O/E, base / + AAC** |
| --- | --- | --- | --- | --- | --- | --- | --- | --- | --- |
| Primary composite  (n = 3,580; 194 events) | Clinical | 9.3 | 0.002 | 0.685 | 0.693 | 0.007 (-0.001-0.024) | 0.004 (-0.008-0.017) | 0.90 / 0.89 | 0.96 / 0.97 |
|  | Clinical + MFR + TPD | 8.3 | 0.004 | 0.702 | 0.709 | 0.007 (-0.001-0.022) | 0.007 (-0.003-0.018) | 0.84 / 0.85 | 0.97 / 0.98 |
| Heart failure admission  (n = 3,569; 95 events) | Clinical | 8.9 | 0.003 | 0.801 | 0.810 | 0.009 (-0.001-0.027) | 0.010 (-0.004-0.023) | 0.91 / 0.88 | 0.98 / 1.00 |
|  | Clinical + MFR + TPD | 6.8 | 0.009 | 0.809 | 0.816 | 0.007 (-0.001-0.023) | 0.008 (-0.004-0.021) | 0.89 / 0.88 | 0.97 / 1.00 |

**Table S3.** Overview of sensitivity analysis results. Sensitivity was assessed for the full versus the triple-normal cohort, for a triple-normal cohort with relaxed CAC criterion (CAC 0-100 instead of 0), for descending thoracic versus ascending aorta calcification, for stress versus rest and for splitting the cohort at the median AAC as opposed to continuous evaluation. All other parameters were unchanged compared to the primary composite endpoint analysis. AAC=ascending aortic calcification. adjHR=adjusted hazard ratio. CAC=Coronary artery calcium. CI=Confidence interval. CT=computed tomography. uHR=unadjusted hazard ratio.

| **Sensitivity to** | **uHR**  **(95% CI)** | **p value (unadjusted)** | **adjHR**  **(95% CI)** | **p value (adjusted)** |
| --- | --- | --- | --- | --- |
| full cohort | 1.48 (1.43-1.53) | <0.001 | 1.11 (1.07-1.15) | <0.001 |
| CAC 0-100 | 1.38 (1.25-1.53) | <0.001 | 1.23 (1.11-1.37) | <0.001 |
| Thoracic calcification | 1.37 (1.19-1.59) | <0.001 | 1.19 (1.03-1.39) | 0.022 |
| stress CT | 1.45 (1.28-1.64) | <0.001 | 1.26 (1.11-1.43) | <0.001 |
| Median split (AAC>0) | 1.76 (1.34-2.33) | <0.001 | 1.44 (1.08-1.91) | 0.014 |

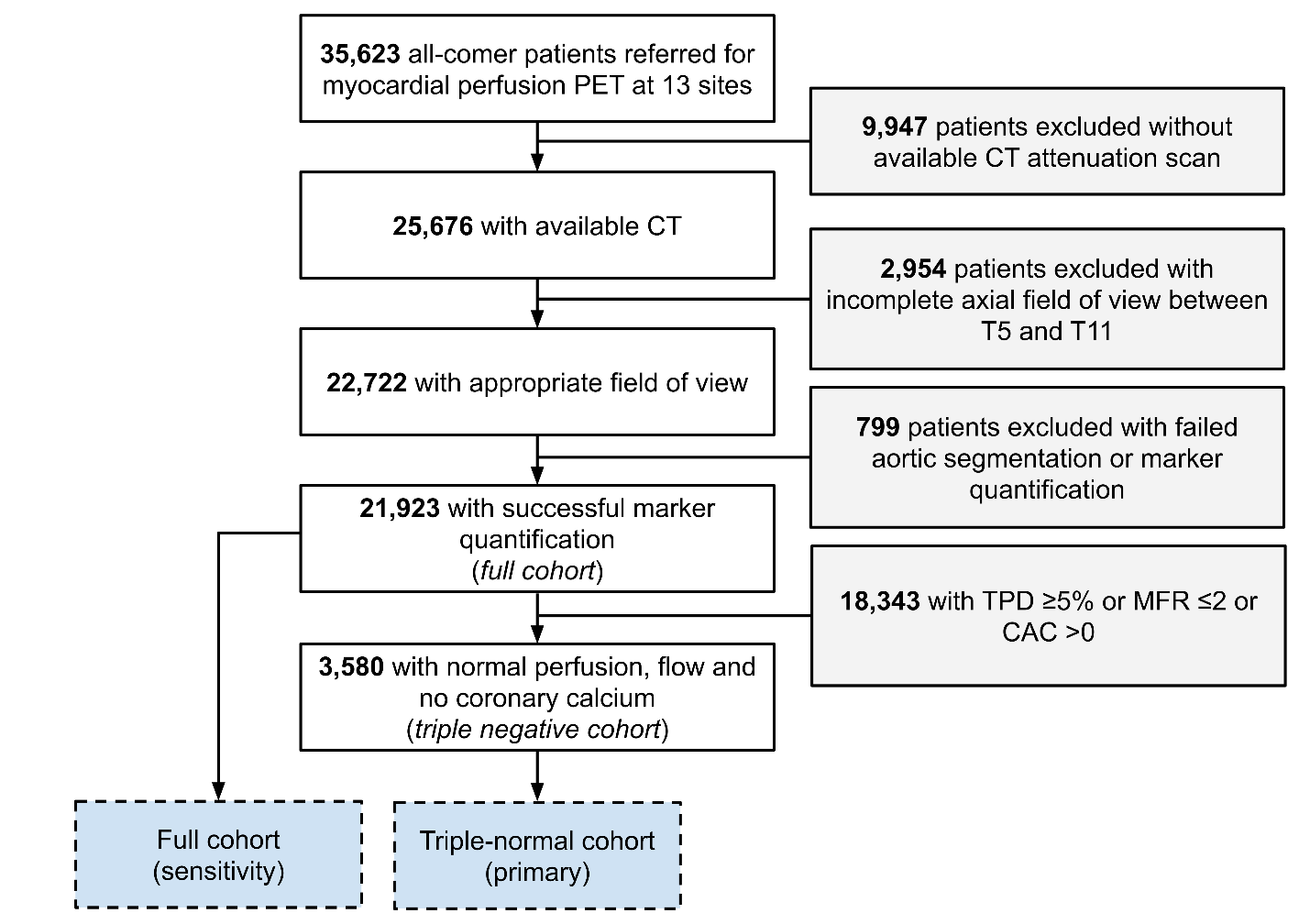

**Figure S1.** Cohort flow diagram. TPD=total perfusion deficit; MFR=myocardial flow reserve; CAC=coronary artery calcium

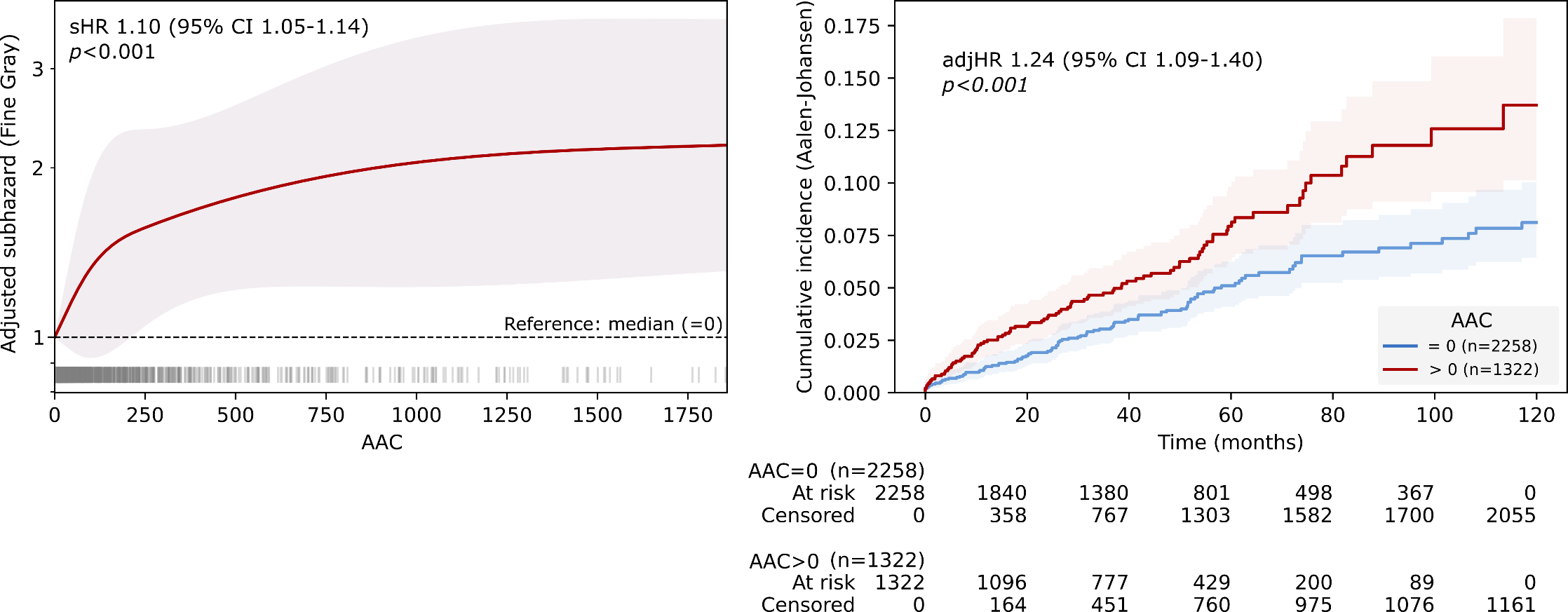

**Figure S2**. Restricted cubic spline analysis for the competing risks model (Fine-Gray) for the composite cardiovascular endpoint (heart failure hopsitalization, cardiovascular death or myocardial infarction). Higher AAC was associated with increased hazard. sHR=subhazard ratio; AAC=ascending aortic calcification

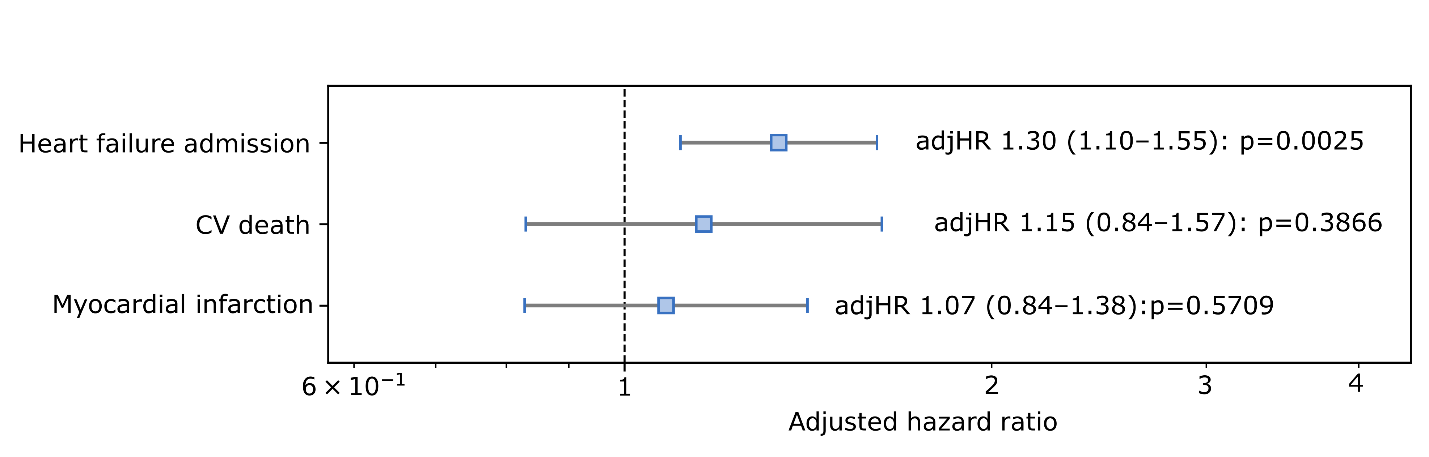

**Figure S3**. Adjusted hazard ratios for the individual components of the composite cardiovascular endpoint. The strongest association was found for heart failure admission. Point estimates were concordant for all three component endpoints but did not reach significance for cardiovascular death and myocardial infarction. adjHR=adjusted hazard ratio; CV=cardiovascular
